# Functional Brain Networks in Focal Dystonia and their Associations with Dystonic Behavior

**DOI:** 10.1101/2021.05.14.21257239

**Authors:** Noreen Bukhari-Parlakturk, Michael Fei, James Voyvodic, Simon Davis, Andrew Michael

**Author notes:** emails addresses. **Declaration of interest:** The authors report no relevant disclosures. **Data availability statement:** Data can be made available after approval from Duke School of Medicine’s Institutional Review Board. The codes can be made available upon a reasonable request to the corresponding author. **Ethics approval, Patient consent statement, and Clinical trial registration:** The human participants participating in the functional MRI experiment and behavioral testing were of a dystonia study. The study was approved by Duke University Health System Institutional Review Board (#0094131), ensuring ethical guidelines were followed and all participants provided written informed consent to participate according to the Declaration of Helsinki. The study was not registered as a clinical trial because it was a small exploratory study. **Authorship statement**: NBP was involved in all aspects of the research project, from conceptualizing the study to data collection, analysis, and manuscript writing. MF was involved in data analysis and assisted with manuscript writing. JV assisted with the study design, data collection and reviewed the manuscript. SD assisted with the study design, provided input on data analysis and feedback on the manuscript, AM helped conceptualize the research study, performed and critiqued data analysis, and assisted with manuscript writing.

## Abstract

Multiple neuroimaging studies suggest that dystonia is a network-level brain disorder, but the key networks to target for brain therapy in dystonia remain poorly understood. This study identified impaired functional networks (FNs) in writer’s cramp (WC) dystonia while participants performed hand motor tasks in the scanner. Task-fMRI images were acquired from twelve WC and twelve healthy volunteers (HV) while they performed writing, tapping, and flexion-extension motor tasks. Group independent component analysis was used to derive FNs, and functional network connectivity (FNC) between the FNs was computed to evaluate their associations with dystonic behavior. Our approach revealed three novel findings: First, we found that the basal ganglia, orbitofrontal, and superior parietal FNs were aberrant during the on-block of the task and, interestingly, also during the off-block period reminiscent of the loss of network inhibitory response during writing in WC. Second, we found multiple impaired FNC patterns in WC primarily in these three FNs these results were further validated through non-parametric testing via Monte Carlo simulations. Third, using FNC deficits and FNC-writing legibility correlations, we characterized these FNC patterns as primary or secondary associations with the motor program for writing. Findings from our data-driven whole-brain analysis approach reveal new FNs disrupted in dystonia and they may serve as key targets for the treatment of this disorder.

**Impact Statement:** Dystonia is a rare and disabling brain disorder affecting motor function. In this study, novel functional network differences in dystonia were identified. Associations between functional network connectivity and dystonic behavior were found. Through a data-driven approach, this study reveals new targets for dystonia treatment.

## INTRODUCTION

Dystonia is a rare and disabling brain disorder characterized by abnormal and painful postures that can manifest over the whole body (generalized dystonia) or isolated in a single limb (focal dystonia) (Albanese et al. 2013). The isolated focal dystonias have a prevalence of 15 per million persons (Warner et al. 2000; Ortiz et al. 2018). A subset of focal dystonias manifest symptoms only during a specific motor behavior. Writer’s cramp (WC), for example, is a task-specific focal dystonia, with symptoms occurring specifically during the task of writing (Albanese et al. 2013; Warner et al. 2000). WC dystonia is an ideal subtype to identify the neural underpinnings of dystonia as the dysfunctional tasks (e.g., writing) can be used to study brain activity using functional magnetic resonance imaging (fMRI) (Hallett 2006; Goldman 2015).

Prior fMRI studies of focal dystonia patients suggest that dystonia is a network-level brain disorder with abnormalities reported during both rest-fMRI (when participants were not engaged in any task) and task-fMRI (while participants performed the dystonic task). A rest-fMRI study demonstrated decreased connectivity of the premotor and superior parietal regions in the hemisphere contralateral to the affected hand (Delnooz et al. 2012). Another rest-fMRI study showed increased primary sensorimotor connectivity in the left premotor-parietal network (Mantel et al. 2018). This study also reported reduced inter-network (e.g., between network) connectivity between the basal ganglia and cerebellar networks in WC (Mantel et al. 2018).

Together, these rest-fMRI studies indicate that patients with focal dystonia have multiple functional abnormalities both within and between networks. Although, these findings identify the neurobiological underpinnings of dystonia, these abnormalities were observed when the participants were not performing a dystonic task, as such it is unclear if they represent a primary causative change such as an intrinsic brain vulnerability or a secondary compensatory change.

In contrast, task-fMRI studies of WC can identify functional abnormalities in the motor program for writing. One study of WC performing the dystonic writing task with the affected hand reported decreased connectivity between the parietal and premotor brain regions (Gallea et al. 2016), while two other studies reported decreased recruitment of the basal ganglia (Zeuner et al. 2015; Berman et al. 2013). Although, task-fMRI studies of WC have thus far identified abnormalities in functional brain regions, no study to date has performed a network-level analysis of WC during task-fMRI. As a result, it remains unclear which functional networks (FN) are abnormal during the motor program of writing, and the primary/secondary causative associations between these FNs are unknown. Understanding the hierarchy of abnormal FNs and their interactions in dystonia pathophysiology may allow us to develop better targets for brain therapy.

To determine if a FN abnormality represents a primary or secondary brain change, we need to understand the relationship between connectivity deficits and the clinical features of dystonia. A previous study calculated connectivity between the inferior parietal cortex and ventral premotor area and demonstrated an inverse association with WC symptom duration (Gallea et al. 2016). These findings suggest that the connectivity between the parietal and premotor regions may be an important marker of the clinical manifestation and therefore represent a primary defect in WC dystonia. Similar studies in blepharospasm and cervical dystonia have also demonstrated an inverse correlation between measures of disease (e.g., disease duration and symptom severity) and decreased regional brain activity (e.g., superior frontal gyrus and somatosensory cortex) (Huang et al. 2017; Burciu et al. 2017). However, more studies are needed at the FN level in WC to understand the relationship between FN abnormalities and clinical measures of the disease. Understanding these fundamental brain-behavior relationships will, in turn, allow us to distinguish between primary and secondary FN abnormalities with the goal of identifying the primary FN hubs to target for treatment of WC dystonia.

The first aim of this study was to identify FN differences between WC participants and healthy volunteers (HV) while participants performed the dystonic writing task in the scanner. We investigate if the FN differences were unique to just the writing task or also common to two other control motor tasks. The second aim of this study was to understand the relationship between changes in FNC and dystonic behavior. To achieve these aims, we used a data-driven approach that allowed unbiased whole-brain analysis to identify FNs of interest. The FNs were identified during the performance of the three motor tasks: a writing task that elicits dystonia in WC participants and two control motor tasks (complex four-element sequence tapping and simple finger flexion-extension). All motor tasks were performed with the participants’ right hand. FNs were then compared with the timing of each motor task presentation to identify key FNs impaired during the active tasks in WC. These impaired FNs were then evaluated for group differences. FNC was then evaluated to identify inter-network changes during writing. Lastly, we examined if the changes in FNC during writing were linked to a key behavioral measure of writing.

## MATERIALS AND METHODS

### Participants

The study was approved by the Duke Health Institutional Review Board and the work described has been carried out in accordance with the declaration of Helsinki for experiments involving humans. The study was open to all WC patients with dystonia. All enrolled patients were more than three months from the last botulinum toxin injections and were not on oral medication of trihexyphenidyl, a drug used for symptomatic treatment for dystonia. All enrolled WC were diagnosed with isolated right-hand dystonia by a Movement Disorder Specialist through standard medical history and neurological examination. Aged-matched HVs who were right-hand dominant with no structural brain disorders or psychiatric illnesses were also recruited for the study. All enrolled participants gave informed consent and had no contraindication to MRI. A total of fourteen WC and nineteen HV participants were consented and completed fMRI scans. After data collection, one WC participant’s data was excluded due to misdiagnosis of essential tremor. Eight additional participants were excluded due to extensive head motion in the fMRI as detailed under the Data Acquisition and Preprocessing section. All analysis and results of this study are based on twelve WC and twelve HV participants.

### Dystonia Behavioral Analysis

At the initial screening visit, all study participants were asked to write a single sentence ten times in MovAlyzer (Neuroscript) software and four times in OneNote (Microsoft) software on a digital tablet. Participants were then asked to perform a twenty-minute writing exercise to induce writing dystonia, then repeat the single sentence using the same software. Writing samples performed at baseline and after the twenty-minute writing exercise were then analyzed for the number of dysfluent events during writing using the MovAlyzer software. Writing samples were also analyzed for legibility using the writing to text conversion feature in OneNote at baseline and after the twenty-minute writing exercise. The total number of words correctly converted from the participant’s writing to a text format across the four sentences was calculated as a percentage of a total of 36 words. A previous study by our group has shown that these writing software measures were comparable to established dystonia rating scales, can capture the significant differences between HV and WC participants, and allow us to use them as reliable and objective measures for clinical characterization of WC dystonia (Bukhari-Parlakturk et al. 2021).

### fMRI Research Design

All participants completed a single fMRI scan session during which they were presented with visual instructions to perform the three motor tasks on an MRI-compatible digital tablet. The three motor tasks were performed with digits of the right hand and consisted of 1) sentence writing (hereafter referred to as “writing”), 2) a four-digit sequence tapping (hereafter referred to as “tapping”), and 3) performing finger flexion and extension movements (hereafter referred to as, “extend”). During the writing task, participants were shown a sentence to copy. During the tapping task, participants were shown a four-number sequence every four seconds and asked to type it out on a four-button keypad. In the extend task, participants were presented with an alternating open and close image of the fingers for 0.4 and 0.6 seconds, respectively, and were asked to match their finger movements with the image. The tapping and extend tasks were selected as control tasks to compare against the writing task as they did not elicit dystonia and surveyed different levels of motor complexity. Tasks were presented in a block design: twenty seconds of task (on-block) alternated with sixteen seconds of rest (off-block) (**Figure 1**). Twelve on-blocks and thirteen off-blocks were presented in a single run lasting seven minutes and twenty-seven seconds for each of the three tasks. The task presentation timing was kept the same across tasks and participants.

**Figure 1:**
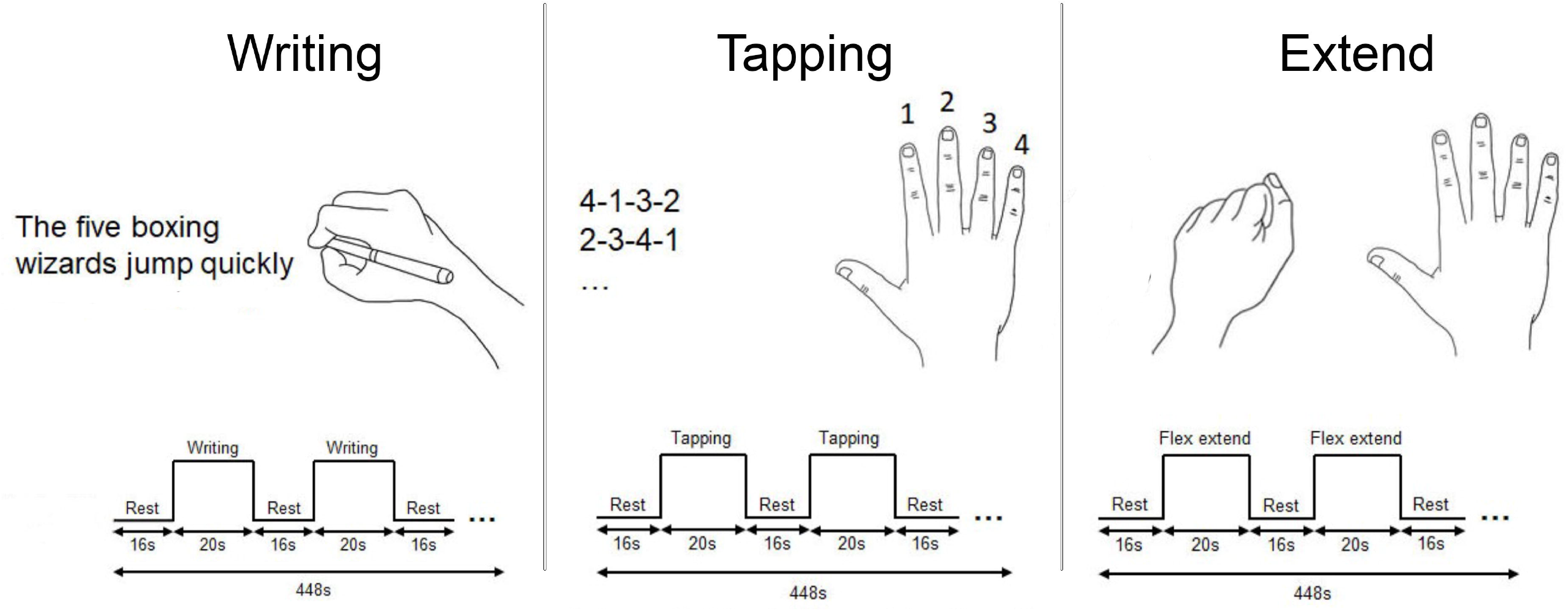
Schematic Illustration of the fMRI Paradigm for the Writing, Tapping and Extend Tasks. Each motor task was presented in a block design for twenty seconds (on-block) alternated by sixteen seconds of rest (off-block). Twelve blocks were presented for each task. Subjects performed all tasks with their right hand. **Writing task:** subjects were presented a holo-alphabetic sentence to copy on a digital tablet. **Tapping:** subjects were asked to tap a four-digit sequence presented every four seconds in a twenty second block. **Extend:** subjects were asked to mimic an image of right hand opening and closing.

### Data Acquisition and Preprocessing

Structural and functional MRI were acquired from all participants using a 3 Tesla GE scanner. Structural T1-weighted images were acquired using echo-planar imaging with the following parameters: voxel size = 1×1×1.3 mm^3^, TR = 6.836 s; TE = 2.976 s; FOV= 25.6mm^2^, bandwidth 41.67 Hz/Pixel. Functional echo-planar images were acquired while participants performed one of the three motor tasks using the following parameters: voxel size = 3.5×3.5×4.0 mm^3^, TR = 2 s, TE = 30 ms, flip angle = 90°, FOV = 22 cm, bandwidth = 250 Hz/Pixel. During the functional scans, participants were asked to minimize their head movement. The CIGAL software was used to present task cues and record and display each participant’s finger movements in real-time (Voyvodic 1999). All fMRI images were preprocessed using fMRIPrep, an automated pipeline to perform brain extraction, head motion, distortion and slice timing correction, intraparticipant registration, and spatial normalization (Esteban et al. 2019). fMRIPrep also provided an estimation of scan quality for each fMRI run. Data from one WC and seven HV were excluded due to excessive head movement as defined by mean frame-wise displacement > 0.5mm.

### Group Independent Component Analysis (GICA)

A GICA analysis was performed on all fMRI scans that passed the motion threshold using the GIFT v3.0 software on MATLAB R2018b (Calhoun et al. 2001). GICA uses a data-driven approach to identify the group-level spatially independent FNs and then projects them onto individual fMRI data to identify FNs at the individual level. GICA also constructs the time courses for each FN at the individual level. GICA was applied to fMRI data across all three motor tasks to identify common FNs using the InfoMax algorithm. GICA was run twice to decompose the group fMRI dataset into 20 and 30 FNs. FNs were then visually inspected and labeled by comparing them against the independent component network atlas software (Smith et al. 2009; Laird et al. 2011; Ray et al. 2013; Kozák et al. 2017) and using the brain regions defined by the Harvard Oxford MNI atlas (Desikan et al. 2006). Compared to the 20-component analysis, spatial maps of the FNs obtained from the 30-component analysis better matched with atlas FNs and all further analyses were based on FNs from the 30-component analysis.

Seven components that primarily corresponded to the cerebrospinal fluid, non-brain tissue and those related to motion artifacts were excluded from further analysis. The remaining components showed a high anatomical overlap with FNs identified in previous studies, indicating independent validation and biological relevance of our components. FNs previously shown to play a role in patients with dystonia (e.g., basal ganglia, cerebellum, superior parietal, sensorimotor, visual, orbitofrontal, and default mode) (Battistella and Simonyan 2019; Mantel et al. 2018; Huang et al. 2017; Baltazar et al. 2020) were then selected for further analysis. The sensorimotor FN was lateralized, and both components were selected for further analysis.

### Correlation of FN Activation to Task Presentation

To identify the FNs that were modulated by the motor tasks, the task presentation time course was convolved with the hemodynamic response function (HRF) to construct a task model time course for all three tasks. The model time courses were then correlated with the time series of the FNs that were estimated by GICA. These correlations between the task model time courses and each participant’s FN time series were compared between the HV and WC groups. Note that this is the only instance in our analyses where we apply the HRF, and this step was performed to validate the temporal relevance of the FNs to the three motor tasks performed. By this step, we also show how GICA can uncover FNs that are closely related to task presentation using a completely data-driven approach.

### FN Average Time Course

Time courses from GICA of the FNs of interest during the writing task were averaged over the writing task blocks to derive participant-specific hemodynamic response for the on-block and off-blocks. With this approach, we aim to investigate the potential timing delays between WC and HV.

### Functional Network Connectivity (FNC)

FNC measures were determined by computing pairwise correlations between the FN time courses. FNC matrices were constructed for FNs of interest for each participant (Jafri et al. 2008). FNC differences between HV and WC were also computed and presented as two-sample t-test values where positive t-values correspond to decreased FNC in WC compared to HV, and negative t-values correspond to increased FNC in WC.

### Association of FNC with Writing Legibility

Aberrant FNCs in WC during writing were then correlated with the behavioral measurement of writing legibility.

### Statistical Analysis

Group-level differences in fMRI mean framewise displacement parameter and age of participants were tested using an unpaired two-sample t-test. Gender differences between the two groups were tested by Fisher’s exact test. Correlation of FNs to the motor task time courses was performed using Pearson’s correlation. FN correlation to the motor task time course was tested between the groups using a two-sample t-test. We further validated the results of the FN correlation to the motor task time course through non-parametric testing using Monte Carlo simulations. Within each simulation we randomly assign pseudo labels to the 12 HV and 12 WC subjects. We repeated this random assignment 10,000 times to create a null distribution for each FN correlation to the motor task time course between pseudo-WC and pseudo-HV. Using the mean and standard deviation of the null distribution we compute the statistical significance of the FN correlation difference between true-WC and true-HV which is presented as a z-score (Hastie, T., Tibshirani, R., Friedman, J.H. and Friedman 2009). For FN hemodynamic response time courses, a two-way repeated-measures ANOVA was performed with factor 1 denoting group (HV vs. WC) and factor 2 time. Interactions between group and time were then analyzed using the *post hoc* Holm Sidak test to determine if there was a significant difference in FN time courses between WC and HV. For FNC analysis, Pearson’s correlation coefficient was transformed to a Fisher’s z-score and compared between the two groups using a two-sample t-test. We further validated the FNC results through non-parametric testing using Monte Carlo simulations as described above (Hastie, T., Tibshirani, R., Friedman, J.H. and Friedman 2009). Key FNCs that showed changes in WC were then correlated at the group level with the behavioral measure of writing legibility using Pearson’s correlation. To determine if there are group-level differences in correlation between writing legibility and FNC between left and right sensorimotor networks, a full factorial general linear modeling was performed after adjusting for age.

## RESULTS

### Participants

We present results from 12 WC and 12 age-matched HVs that passed motion threshold standards. Participant demographics, head motion, and writing behavioral measures are presented in **Table 1**. There were no significant group differences in age or gender. WC showed a mean disease duration of 9.6 years (SD 14.5). There were no group differences in head motion during the three runs of motor task-fMRI (writing p=0.35, tapping p=0.66 and extend p-0.40). Outside the fMRI scanner, analysis of writing samples from WC showed significantly higher dysfluent events (p=0.009) and decreased legibility (p=0.037) relative to HV, providing a quantitative measurement of their WC dystonia severity (**Table 1, Writing Behavioral Measures**). These results indicate that the dystonic behavior measured in our patient cohort occurred during the writing task.

**Table 1:** Subject Demographics, fMRI Head Motion and Behavioral Measures.

|  | WC (n = 12) | HV (n = 12) | WC vs HC<br>t-test, p-value |
| --- | --- | --- | --- |
| **Demographics** |  |  |  |
| Age (yrs) | 54.8 (13.7) | 54.1 (11.9) | 0.90 |
| Gender (F/M) | 3/9 | 6/6 | 0.40 |
| Disease<br>Duration (yrs) | 9.6 (14.5) | N/A |  |
| **fMRI Mean Framewise Displacement (mm)** |  |  |  |
| Writing | 0.22 (0.11) | 0.18 (0.07) | 0.35 |
| Tapping | 0.15 (0.07) | 0.14 (0.05) | 0.66 |
| Extend | 0.20 (0.11) | 0.17 (0.05) | 0.40 |
| **Writing Behavioral Measures** |  |  |  |
| Dysfluent<br>events (#) | 439 (182) | 284 (48) | 0.009** |
| Writing<br>Legibility (%) | 46 (29) | 73 (29) | 0.037* |

Data presented as values presented are mean (SD). \*p<0.05, and \*\*p<0.01

### Functional Networks

Visual inspection of the FNs revealed that eight of the 30 FNs in our ICA analysis closely resembled previously reported FNs that used rest-fMRI (Smith et al. 2009) and task-fMRI on independent samples (Laird et al. 2011; Ray et al. 2013; Kozák et al. 2017). Specifically, we report FNs for the left and right sensorimotor network (SMN-L, SMN-R, Laird ICN 8), visual network (VISN, Laird ICN 11), cerebellum network (CBLN, Laird ICN 14), basal ganglia network (BGN, Laird ICN 4), superior parietal network (SPN, Laird ICN 7), orbitofrontal network (OFCN, Laird ICN 2), and default mode network (DMN, Laird ICN 13). The BGN, CBLN, VISN reported in this task-fMRI analysis also resembled prior resting-state fMRI FNs of WC participants (Mantel et al. 2018). However, our task-fMRI study showed a different cortical network composition than Mantel and colleagues (**Figure 2 and Table 2**). Specifically, the SMN did not divide into a dorsal and ventral network but rather laterally, with greater left and greater right activated FN and can be attributed to the fact that participants performed motor tasks with the right hand.

**Figure 2:**
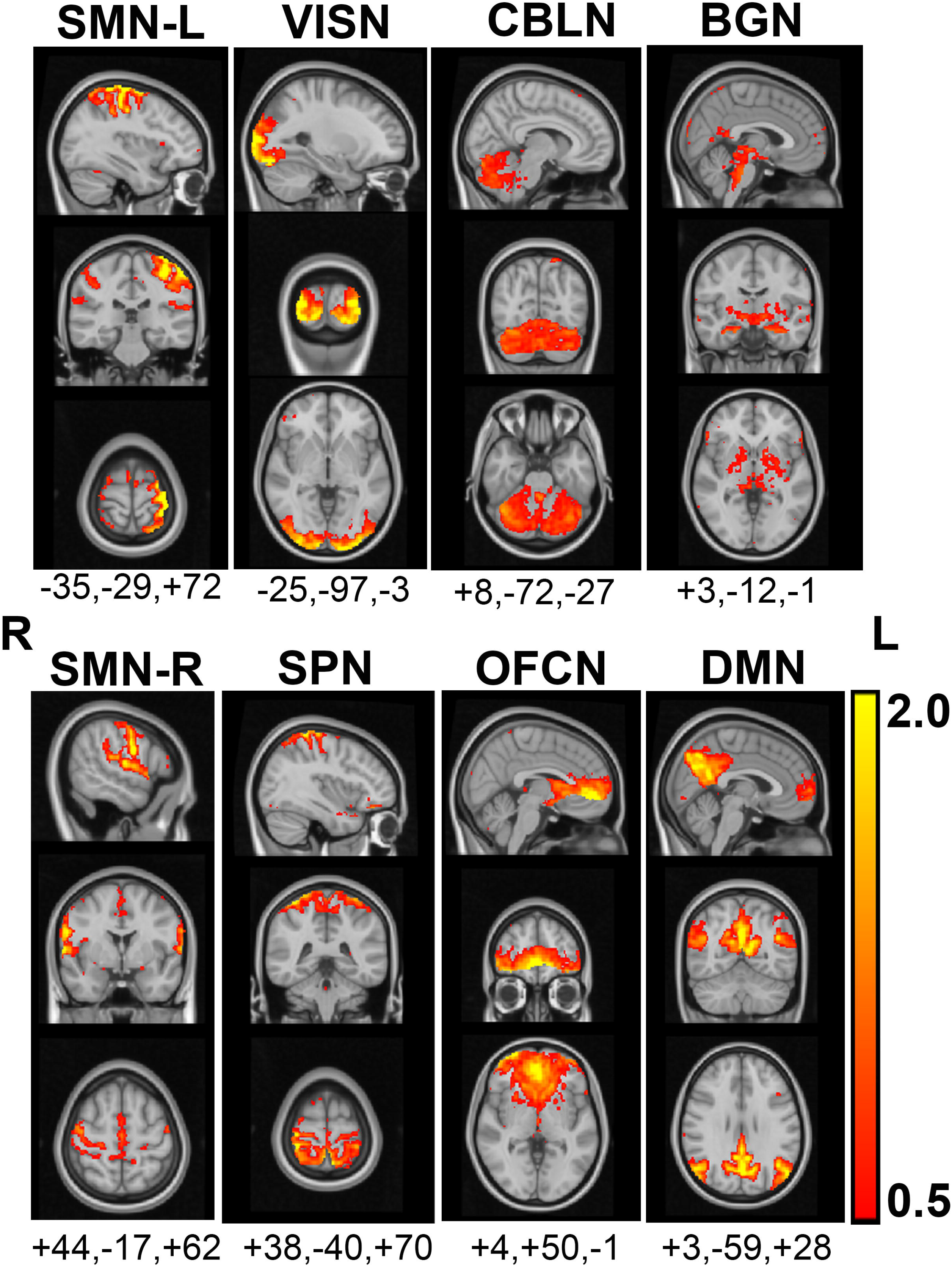
Functional Networks (FNs) derived from task fMRI of the three motor tasks. Anatomical details of the FNs are provided in Table 2. FNs are shown in sagittal, coronal and axial planes at peak activation with MNI coordinates (displayed in radiological convention).

**Table 2:** Functional Networks identified through the data-driven approach and their anatomical regions.

| Functional network | Description | Anatomical regions* |
| --- | --- | --- |
| SMN-L | Left sensorimotor network | Left precentral gyrus, left postcentral gyrus, right postcentral gyrus |
| VISN | Visual network | Bilateral occipital cortices |
| CBLN | Cerebellum network | Cerebellum (bilateral lobules and vermis) |
| BGN | Basal ganglia network | Bilateral basal ganglia (putamen, globus pallidus), thalami, bilateral temporal cortices, brain stem |
| SMN-R | Right sensorimotor network | Right precentral gyrus, right postcentral gyrus supplementary motor cortex, left postcentral gyrus |
| SPN | Superior parietal network | Bilateral superior parietal lobes |
| OFCN | Orbitofrontal network | Bilateral frontal poles, bilateral caudate, paracingulate gyrus |
| DMN | Default Mode Network | Precuneus, cingulate gyrus (posterior division), bilateral angular gyrus, superior frontal gyrus, lateral occipital poles |

\* In the descending order of cluster size

### Correlation of FN Activation to Task Presentation

For each group, we evaluated the strength of the correlations of FN time courses to the timing of task presentation and ordered the FNs according to descending strength of correlation **(Figure 3)**. Our analysis revealed that WC (red) activated the same FNs during writing as their HV counterparts (blue).

**Figure 3:**
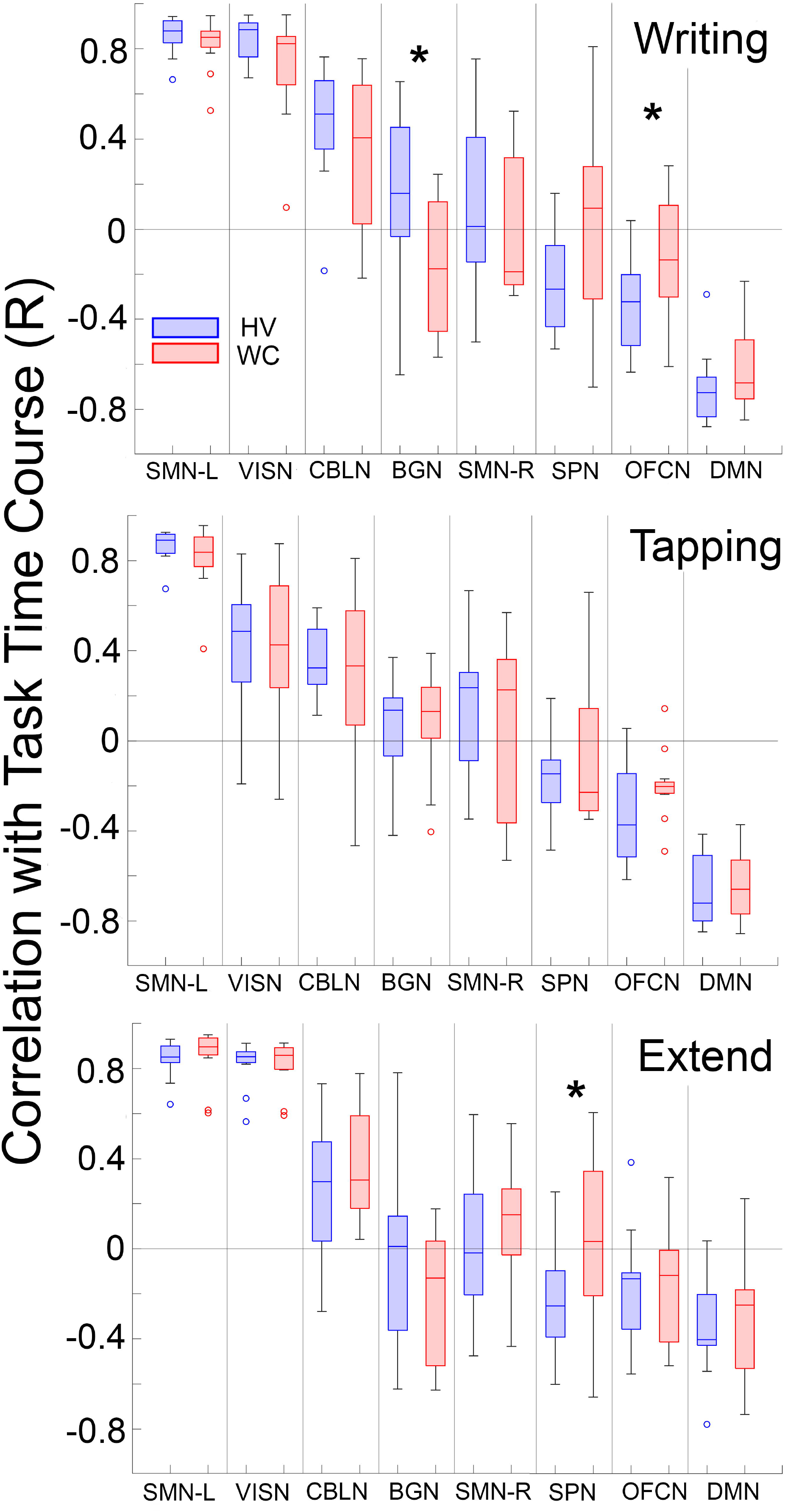
Correlation of Functional Network Activation to Motor Task Presentation. The functional networks (FNs) are ranked from highest to lowest correlation for the writing task and FNs are presented in the same order for tapping and extend tasks. Box plot represents 95% of variability across subjects and whiskers represent the interquartile range with outliers shown. * denotes significant (p<0.05) difference between HV and WC.

We next examined how these FN relationships manifested in WC. Compared to HV, most FNs in WC showed a weaker median correlation with the writing task **(Figure 3, top graph)**. Notably, this trend for weaker correlation strength in WC was not observed in the tapping or extend tasks **(Figure 3, middle and bottom graphs respectively)**. Group differences of each of the FNs are discussed below. We further validated group differences in FNs with a non-parametric approach **(Table 3)**.

**Table 3:** WC and HV Show Significant Group Difference in FN Correlation with Task Time Course.

| Task | FN | HV vs WC |  | MC |  |
| --- | --- | --- | --- | --- | --- |
|  |  | t-value | p-value | z-score | p-value |
| Writing | BGN | 2.60 | 0.02* | -2.31 | 0.02 |
| Writing | SPN | -1.2 | 0.07 | 1.82 | 0.07 |
| Writing | OFCN | -2.25 | 0.03* | 2.07 | 0.04 |
| Extend | SPN | -2.15 | 0.04* | 2.02 | 0.04 |

Note: Group comparison (HV vs WC) of task-correlation values for each FN are provided as a critical t-value and p-value for two tailed t-test. MC z-score and p-value represent results of Monte Carlo randomized group simulation with 10,000 permutations. \*p<0.05

#### *Left Sensorimotor Network (SMN-L*)

The SMN-L showed the strongest positive correlation (>0.8) to all three tasks. This finding was expected as the three tasks are motor in nature and performed with the right hand. The strong correlation between the SMN-L and the task time course serves as a positive control indicating that FNs produced by GICA are valid both in terms of the anatomical relevance and association with task presentation. The SMN-L showed the lowest intra-group variance across all three tasks in both HV and WC groups. There was no significant difference between HV and WC at the group level for the three motor tasks. These results indicate that the SMN-L was modulating very closely to task presentation in both groups across all participants.

#### Default Mode Network (DMN)

The DMN showed the most negative correlation to all three task time courses in both groups. This result further validates the FNs obtained through GICA as DMN is expected to negatively correlate to task presentation. At the group level, there was no difference in the correlation of the DMN to the three motor tasks between HV and WC.

#### Basal Ganglia Network (BGN)

The BGN correlated with the writing task time course, but the direction of correlation varied by group. In HV, the BGN was positively correlated with the writing task time course. However, in WC, the BGN was negatively correlated and was significantly different from HV (p=0.02). For the tapping task, the BGN in both groups was slightly positively correlated with the task time course, but no group difference was observed. For the extend task, HV showed no correlation to the task time course, while the WC group was slightly negatively correlated with no significant group difference.

#### Orbitofrontal Network (OFCN)

The OFCN negatively correlated with all three task time courses in both HV and WC. However, in the writing task, WC showed a lower negative correlation to task timecourse compared to HV, and this difference was statistically significant (p=0.03). No significant group differences were observed for the tapping and extend task time courses.

#### Superior Parietal Network (SPN)

The SPN negatively correlated with all three motor task time courses in HV. For the extend task, WC participants showed a more negative correlation to the task than HV, and this difference was statistically significant (p=0.04). For the writing task, although a trend of significant group difference was observed, the group difference was not statistically significant (p=0.07). The lack of statistical significance can be attributed to the high variability observed in the WC.

#### Cerebellum Network (CBLN)

The CBLN positively correlated with all three task time courses. No significant group-level differences were observed between WC and HV. However, a higher intra-group variance was observed in WC to the writing task.

#### Right Sensorimotor Network (SMN-R)

The SMN-R showed no correlation with HV participants’ writing and extend task time courses. In contrast, in WC, the SMN-R negatively correlated with the writing task time course and positively correlated with the extend task time course. The SMN-R showed a positive correlation with the tapping task time course in both groups.

To further validate group differences in the correlations of FN time courses to the timing of task between WC and HV during the 3 tasks, we ran a non-parametric Monte Carlo simulation (10,000 permutations) that randomized subjects across the two groups **(Table 3**). The difference in FN time courses between true-HV and true-WC was significantly different from difference of randomly generated groups (e.g. z-score = -2.31, p =0.02 for BGN during writing). In summary, comparisons of WC vs. HV FN time courses with task presentation showed the most significant differences in the BGN, OFCN, and SPN. The BGN and OFCN are distinguished by showing deficits selective to the writing motor task and not tapping or extend tasks.

### FN Average Time Courses

We then analyzed the time course of activity of the three FNs (BGN, OFCN, SPN) that showed disrupted correlation with the motor task time courses in WC presented in the previous section. As a control, we compared these three FNs to the SMN-L, which showed no group differences in correlation with the motor task time courses. We subdivided all four FNs into the on-block and off-blocks during writing to determine where differences in activation occurred (**Figure 4**). As expected, the SMN-L showed no group differences during the on-block or off-block (HV vs. WC, p=0.592). By contrast, the BGN in WC showed a significant delay in peak activation during the on-block (HV vs. WC at 6-10s and 16s, p<0.05) and did not return to baseline during the off-block compared to HV (HV vs. WC at 28-36s, p<0.05). In SPN, WC and HV showed activations in opposite directions; HV showed negative activation during on-block (HV vs. WC at 8-12s, p<0.05), and WC showed negative activation during off-block (HV vs. WC at 32-36s, p<0.05). Lastly, in the OFCN, although the activation in both groups followed the same pattern, i.e., negative correlation to task presentation during on-block, the WC was slower in getting back to the baseline during the off-block (HV vs. WC at 20s, p <0.05; HV vs. WC at 22s, 32s, and 36s, p<0.05). Overall, we note that the SMN-L that was highly correlated to the writing task time course and did not show group differences in its activation. Whereas BGN, SPN, and OFCN FNs that were not highly correlated to task time course, showed significant group differences in their activation during both the on-block and off-block periods of writing.

**Figure 4:**
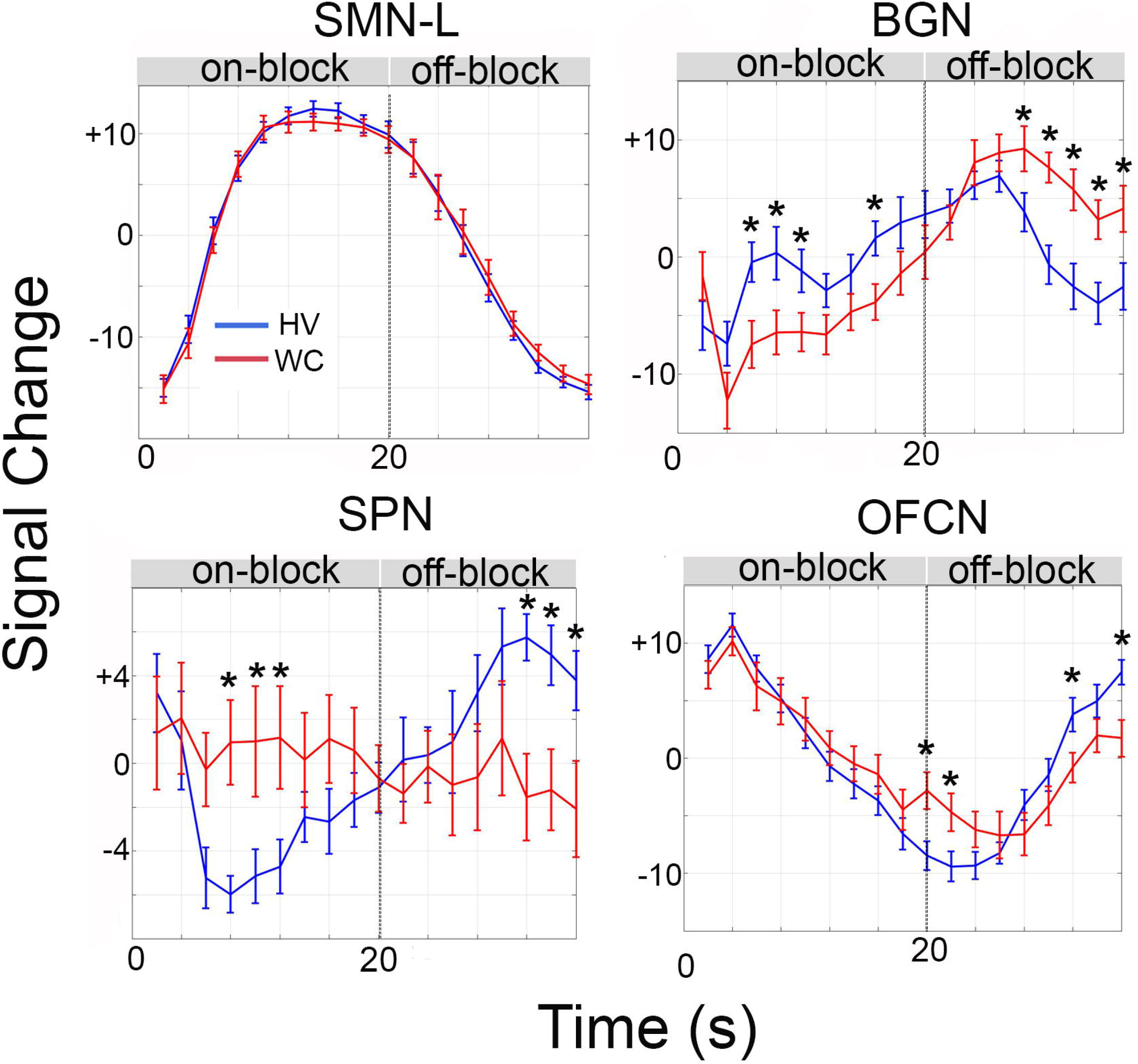
Average Timecourse of Functional Networks (FNs) during writing: FN timecourses averaged across writing task blocks for HV (blue) and WC (red). Dashed line separates on-block and off-block periods. Data shows mean ± SEM. * denotes significant (p<0.05) difference between groups at a specific time and was calculated using two-way repeated measures ANOVA and post-hoc *Holm Sidak* test for interaction between group (HV vs WC) and time (0-36s).

### Functional Network Connectivity (FNC)

FNC differences between the two groups for the writing task are presented in **Figure 5**. In general, we found HV versus WC FNC differences in the BGN, OFCN, and SPN. We further validated these FNC results with a non-parametric approach. We discuss these differences below.

**Figure 5:**
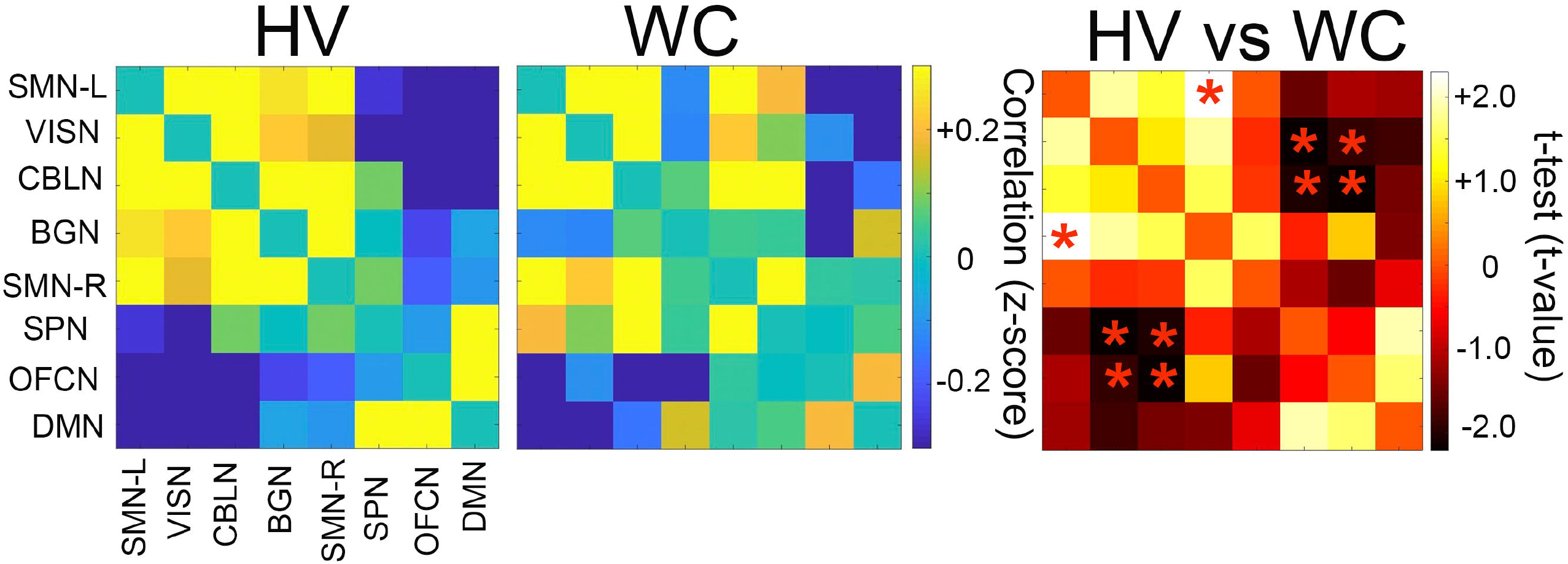
Functional Network Connectivity (FNC) during writing task. Eight FN time courses during writing task was correlated to each other in HV and WC. The FNC values (Fisher’s z-score) are presented as a color matrix with the strength of correlation shown on a blue-yellow color bar. Differences in FNC between HV and WC were evaluated by a t-test. T-values for each FNC is presented in a color matrix with positive t-values corresponding to decreased FNC and negative t-values corresponding to increased FNC in WC compared to HV. * denotes significant (p<0.05, uncorrected) difference between HV and WC.

FNC between BGN and SMN-L was positive in HV but negative in WC, and this difference was statistically significant (**Figure 5, Table 4**). Conversely, strongly negative FNC was noted in OFCN-CBLN in HV, and WC showed weaker negative correlations. WC also showed strongly positive FNC in SPN-CBLN compared to HV. Furthermore, FNC between SPN and VISN was strongly negative in HV but weakly positive in WC. Lastly, HV showed strongly negative FNC in OFCN-VISN, and WC showed weaker negative correlations. A trend for weaker positive FNC between the BGN-CBLN was observed in WC but did not rise to the threshold for significance (HV = +0.36, WC = +0.07, p = 0.12).

**Table 4:** FNC in WC and HV that Show Significant Group Difference During Writing.

| FNC | HV | WC | HV - WC |  | MC |  |
| --- | --- | --- | --- | --- | --- | --- |
|  |  |  | t-value | p-value | z-score | p-value |
| BGN-SMNL | +0.26 (0.42) | -0.11 (0.31) | +2.49 | 0.02 | -2.25 | .02 |
| OFCN-CBLN | -0.74 (0.31) | -0.33 (0.51) | -2.39 | 0.03 | 2.16 | .03 |
| SPN-CBLN | +0.09 (0.19) | +0.41 (0.47) | -2.22 | 0.04 | 2.05 | .04 |
| SPN-VISN | -0.37 (0.35) | +0.10 (0.56) | -2.43 | 0.02 | 2.21 | .02 |
| OFCN-VISN | -0.39 (0.35) | -0.11 (0.33) | -2.07 | 0.05 | 1.94 | .05 |

Note: FNC values are Fisher transformed mean z-scores with standard deviations provided in parenthesis. MC z-score and p-value represent results of Monte Carlo randomized group simulation with 10,000 permutations.

To further validate the FNC difference between WC and HV during the writing task, we ran a non-parametric Monte Carlo simulation (10,000 permutations) that randomized subjects across the two groups **(Table 4)**. The difference in FNC between true-HV and true-WC was significantly different from FNC difference of randomly generated groups (e.g. z-score = -2.25, p =0.02 for BGN-SMNL). In summary, we identified that WC participants show significantly impaired FNC in only the BGN-SMN-L, OFCN-CBLN, and SPN-CBLN during the writing task.

### FNC Correlation to Writing Legibility

FNCs did not correlate with age or disease duration in the WC group (data not shown). We next examined the relationship between FNs that showed impairment in our previous analyses and a behavioral measure of writing legibility. In HV, we found that writing legibility positively correlated with FNC in BGN-SMN-L (R=0.58, p=0.05) and inversely correlated with OFCN-CBLN (R= -0.64, p=0.02). In WC participants, we found the same trends in these FNs, but the associations were much weaker (BGN-SMN-L: R=0.36, p=0.24, and OFCN-CBLN: R=-0.36, p=0.26). No significant correlations were identified between writing legibility and FNC in SPN-CBLN in either HV or WC groups (data not shown).

We further examined the relationship between writing legibility and two additional FNCs that had been previously identified in the rest-fMRI studies: BGN-CBLN and SMN-L-SMN-R (Mantel et al. 2018; Battistella and Simonyan 2019). Both HV and WC showed positive correlations between legibility and BGN-CBLN FNC, though here again, the strength of WC correlation was weaker (HV: R=0.61, p= 0.04; WC: R=0.41, p=0.19). Strikingly, while HV showed strong inverse correlations between writing legibility and SMN-L-SMN-R (R= - 0.64, p=0.02, **Figure 6**), WC participants did not show a significant association between legibility and SMN-L-SMN-R FNC (R=0.07, p=0.83, **Figure 6**). A group level (HV vs. WC) comparison of this legibility to SMN-L-SMN-R FNC correlation showed a significant difference (p=0.03, GLM analysis with adjustment for age, **Figure 6**).

**Figure 6:**
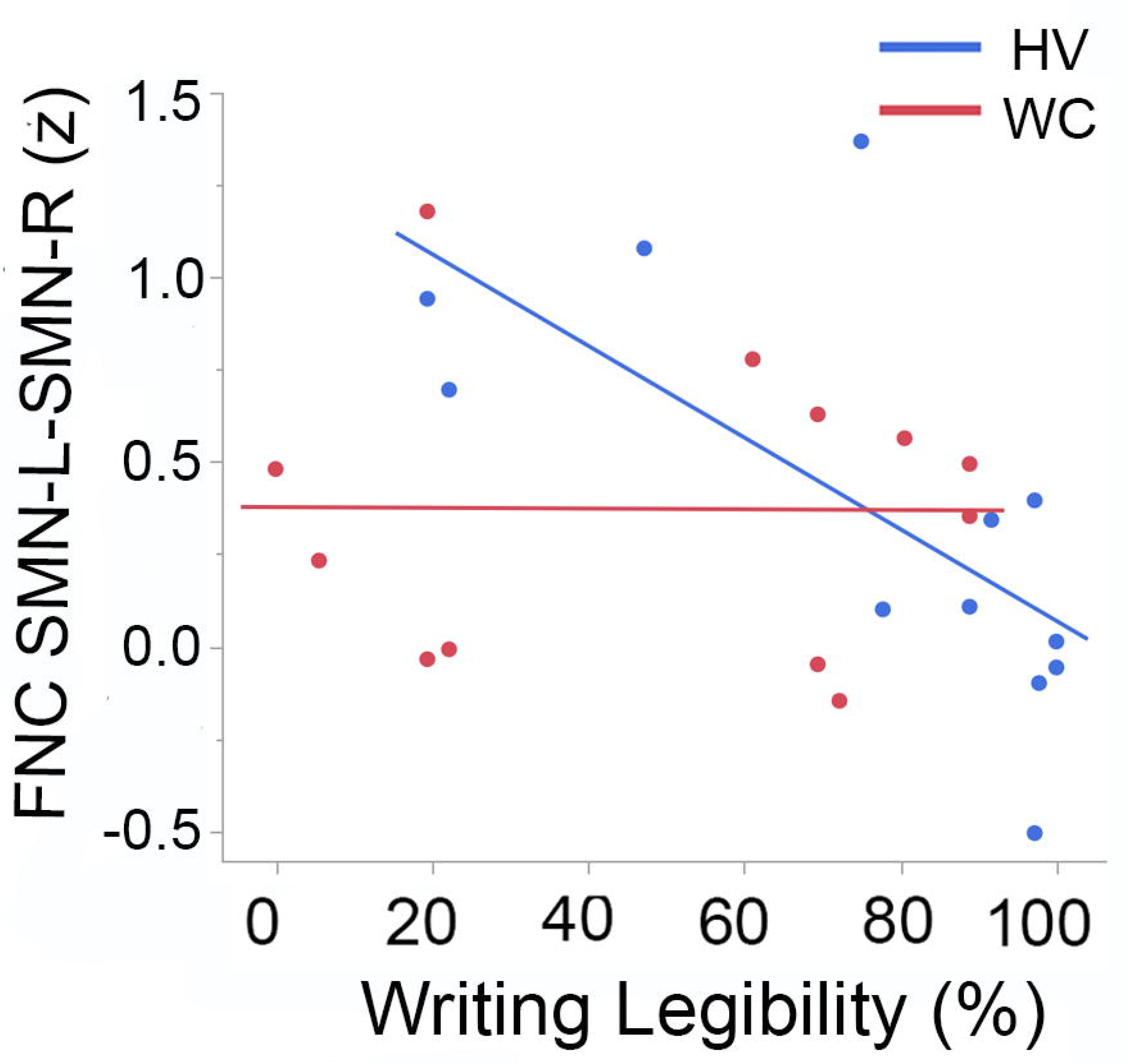
Correlation between functional network connectivity (FNC) and writing legibility. FNC SMN-L-SMN-R during writing task in each HV and WC participant was correlated with the participant’s writing legibility measure. FNC values are Fisher transformed mean z-scores and writing legibility is calculated as percent of correctly written words.

In summary, we find that writing legibility in HV significantly correlates with all four of the examined FNC relationships. WC participants showed weaker trends for these relationships in 3 of the 4 FNC comparisons and uniquely showed a significant group-wise difference from HV in the relationship between legibility and SMN-L-SMN-R FNC.

## DISCUSSION

The aim of this study was to understand the changes in functional networks in writer’s cramp (WC) participants compared to healthy volunteers (HV) during the dystonic writing task. A second aim of this study was to understand the relationship between the functional network (FN) abnormalities and behavioral measures of dystonia. Unlike prior task-fMRI studies, which analyzed regional brain activity, we analyzed abnormalities across multiple functional networks during writing in WC dystonia. Our data-driven whole-brain analysis revealed three novel findings. First, we found that WC showed impairment in three critical FNs: the basal ganglia, orbitofrontal, and superior parietal networks during both on-block and off-block periods of writing, reminiscent of loss of network inhibitory response. Second, we found multiple impaired functional network connectivity (FNC) primarily in these three FNs. Third, drawing on the WC FNC deficits and brain-behavior correlations as well as prior work, we further characterize these FNC patterns as primary or secondary associations with the motor program for writing. Our data-driven whole brain analysis approach thus revealed newly disrupted FNs in dystonia which may serve as key targets for treatment of focal dystonia. We discuss these findings in further detail below.

We report that WC exhibit weaker correlations across three key FNs compared to HV. Using correlations between FN activations and the timing of motor task presentation (**Figure 3 and Table 3**), we demonstrated that the BGN and OFCN were significantly impaired in WC during the writing task and the SPN during the extend task. Thus, the BGN and the OFCN are selectively abnormal in the motor program for writing, while the SPN is abnormal in the general motor program. A follow-up analysis of the time courses of these three FNs during the on-block and off-block of writing revealed that all three FNs showed not only impaired activation during the on-block of writing but also did not return to baseline activity during the off-block period. Our findings are complementary and novel to two prior studies that reported decreased peak activation of basal ganglia and parietal brain regions during the writing and learned tapping task (Zeuner et al. 2015; Gallea et al. 2016). Our findings are complementary to previous findings, as the key brain regions (e.g., basal ganglia, superior parietal region) of each impaired FNs reported in this study have previously been reported as disrupted in the two WC studies. Our findings are novel because we report disruption in multiple simultaneous FNs and FNCs during the writing task with disruptions during both the on-block and off-block periods. We attribute these novel findings to our whole brain ICA analysis approach, which made no assumptions about the hemodynamic response time course, and the networks were identified through a data-driven approach. Our findings also indicate that the FN impairments during the writing task in WC are not common to other motor tasks.

We also report a novel neuroimaging phenotype of persistent FN activation during the off-block of writing (**Figure 4**). This phenotype is reminiscent of the loss of inhibitory response, a principal finding demonstrated in multiple clinical and neurophysiology studies in focal dystonia patients (Hallett 2006; Lozeron et al. 2016). Clinical examples of loss of inhibition are loss of reciprocal inhibition in patients with focal dystonia (Nakashima et al. 1989; Panizza et al. 1990) and loss of blink reflex recovery in blepharospasm patients (Berardelli et al. 1985). Neurophysiology examples of loss of inhibition include loss of short intracortical inhibition using a paired-pulse method and shortened silent periods on voluntary EMG of focal dystonia patients. These abnormal neurophysiological measures reflect a loss of motor cortex inhibition (Hallett 2006). Loss of inhibitory signal was also demonstrated in a PET study of focal hand dystonia patients. Specifically, a PET study using GABA-receptor binding protein showed loss of inhibitory signal in the right cerebellum and left sensorimotor cortex of dystonia patients (Gallea et al. 2018).

Thus, the phenotype of persistently active FNs during the off-block period of writing in WC may be capturing a critical defect in the dystonia pathophysiology, previously only reported in the cortex and cerebellar brain regions in a PET study of human dystonia. Future studies are needed to further characterize this novel neuroimaging phenotype in WC.

Another novel finding of our study is that using WC FNC deficits and brain-behavior correlations, we can hypothesize if a FN abnormality represents a primary or secondary change and thereby begin to construct a hierarchy of abnormal FNs in dystonia to target for brain therapy. In our study, during the select task of writing, WC showed altered FNC with the BGN, OFCN, and SPN. Specifically, we observed decreased FNC between the BGN and SMN-L only during the writing task in WC (**Figure 5 and Table 4**). The brain and behavior correlational analysis showed that in a healthy participant, as writing legibility improves, the connectivity between the BGN and SMN-L increases, while a similar but weaker correlation is found in WC participants. The positive correlation between the FNC and writing legibility suggests that the BGN and SMN-L connectivity may be a primary change in WC. This is consistent with prior functional studies of WC demonstrating decreased cortical-basal ganglia connectivity during writing in task-fMRI (Gallea et al. 2016) and rest-fMRI (Mantel et al. 2018), as well as a structural study showing decreased white matter integrity between premotor cortex and basal ganglia in WC (Berndt et al. 2018). Taken together, findings from our study in conjunction with prior studies suggest that the decreased FNC between the BGN and SMN-L may be a primary change that plays a role in the motor program for writing in WC dystonia.

During the select task of writing, WC also showed altered FN, FNC, and brain-behavior correlation with the OFCN. Specifically, our analysis showed that OFCN negatively correlates to the writing task time presentation in HV (**Figure 3, top graph**), suggesting that the OFCN is modulated during the writing task in HV, and this is impaired in WC. The FNC of OFCN-CBLN is also strongly negative in HV suggesting there’s an inverse relationship between these two FNs, and this is again impaired in WC (**Figure 5 and Table 4**). Taken together, these data suggest that the OFCN-CBLN FNC is potentially an important connectivity pattern for the motor program of writing that has not been previously studied in WC dystonia. Further investigations are needed to understand the contribution of OFCN to dystonia pathophysiology.

Our study did not find FNC differences between the SMN-L and SMN-R. However, based on prior studies, we investigated if the unaffected SMN-R may play a secondary role for the SMN-L abnormalities in WC. Our analysis showed that in HV, as the writing task becomes more demanding (legibility decreases), there is increased FNC between the SMN-L and SMN-R (**Figure 6**), suggesting that the SMN-R is normally recruited with greater task demand. In contrast, as the task demand increases in WC, the FNC between the SMN-L and SMN-R fails to increase. Our findings thus suggest that WC are unable to recruit the SMN-R with increasing task demand compared to their HV counterparts. Taken together, the lack of group differences in SMN-L-SMN-R FNC combined with the strong brain-behavior correlation suggest that the FNC between the SMN-L and SMN-R may be a secondary change in the motor program for writing during increased task demand.

A limitation of our study is the small sample size which led to certain key observations that showed trends but did not reach statistical significance. Future studies of WC using larger sample sizes are needed to confirm the current study findings. Nevertheless, the sample size used in this rare disease cohort was sufficient to demonstrate key FN abnormalities across the three motor tasks in WC compared to HV. Another limitation of our study is that the data-driven approach cannot differentiate between motor planning and motor execution of a task. Future studies using region of interest analysis can differentiate between the two stages of a motor task performance and provide insight into whether both stages of a motor task are abnormal during the on-block and off-block periods of writing. A theoretical concern in neuroimaging studies of dystonia patients is that the FN changes observed during a dystonic task may reflect differences due to the performance of the movement rather than the disease. If this were true, differences in motor performances with the right hand in HV and WC would lead to group differences in the contralateral SMN-L activation and high intra-group variability in the SMN-L. In fact, across all three motor tasks, we saw no group differences in the correlation of the SMN-L to the motor tasks and the smallest intra-group variability of all FNs. Therefore, differences in FNs demonstrated in our study reflect the disease state and not the performance of the motor movement.

## CONCLUSIONS

Our data-driven analysis of WC during the performance of the three motor tasks with their right hand showed that WC activate the same functional networks (FNs) as HV during motor task performance. We demonstrated that the basal ganglia, orbitofrontal, and superior parietal networks show significantly impaired correlation to motor task presentation during the on-block and off-block period of writing, reminiscent of a loss of network inhibitory response in WC. We also found multiple impaired functional network connectivity (FNC) patterns primarily with these three FNs. Lastly, using deficits in FNC and brain-behavior correlations, we characterized FNC patterns as primary or secondary associations with the motor program for writing. Our approach revealed multiple newly disrupted FNs in WC dystonia which may serve as targets for treatment.

## Data Availability

Data available upon request from the authors

## Abbreviations

(WC): Writer’s cramp
(HV): Healthy volunteers
(FN): Functional network
(FNC): Functional network connectivity

## Acknowledgements

The authors would like to thank Amber Holden, Ashley Pifer and Kelsey Ling, who served as Clinical Research Coordinators for this study, and Mariusz-Derezinski-Choo, assisted with fMRI preprocessing. The authors would also like to thank Dr. Nicole Calakos and Dr. Scott Huettel for their input on data analysis and constructive feedback on the manuscript drafts.

## Notes

**Funding:** This work was supported by grants from Dystonia Medical Research Foundation, Doris Duke Charitable Foundation and the National Institutes of Health, National Center for Advancing Translational Sciences (KL2TR002554, TR001456) and National Institute of Neurological Disorders and Stroke (NS065701, NS116025). The content is solely the responsibility of the authors and does not necessarily represent the official views of the funding agencies.

### Competing Interest Statement

The authors have declared no competing interest.

### Funding Statement

This work was supported by grants to NBP from Duke Clinical and Translational Science Association from National Center for Advancing Translational Science (NCATS, 1KL2TR002554), Duke Clinical Translational Science Institute, Dystonia Medical Research Foundation (Clinical Fellowship Training Program from), and Doris Duke Charitable Foundation (Fund to Retain Clinician Scientist). NBP was also supported by a career development award from the Dystonia Coalition (NS065701, TR001456, NS116025) which is part of the National Institutes of Health (NIH) Rare Disease Clinical Research Network (RDCRN), supported by the Office of Rare Diseases Research (ORDR) at the National Center for Advancing Translational Science (NCATS), and the National Institute of Neurological Diseases and Stroke (NINDS).

### Summary of Updates

Functional networks analyzed in this study were reduced from eleven to eight (Figure 2, Figure 3). Statistical analysis was added to compare the brain signal in healthy and dystonia subjects during the on-block and off period of writing (Figure 4). Functional network connectivity analysis was focused on the writing task only (Figure 5). A correlation between brain and behavior measures of dystonia was also added (Figure 6).

